# Real-World Progression-Free Survival as an Endpoint in Advanced Non-Small-Cell Lung Cancer: Replicating Atezolizumab and Docetaxel Arms of the OAK Trial Using Data Derived From Electronic Health Records

**DOI:** 10.1101/2022.05.02.22274571

**Authors:** Shivani K. Mhatre, Robson J. M. Machado, Thanh G.N. Ton, Huong Trinh, Julien Mazieres, Achim Rittmeyer, Michael T. Bretscher

**Author notes:** **Corresponding author information:** Michael T. Bretscher, F. Hoffmann-La Roche Ltd, Grenzacherstrasse 124, 4070 Basel, Switzerland.

## Abstract

**Background:** Evaluating cancer treatments in real-world data (RWD) requires informative endpoints. Due to non-standardized data collection in RWD, it is unclear if and when common oncology endpoints are approximately equivalent to their clinical trial analogues. This study used RWD to replicate both the atezolizumab and docetaxel arms of the OAK trial. Outcomes using progression-free survival (PFS) derived from abstracted physician’s notes in RWD (rwPFS) were then compared against PFS outcomes derived according to Response Evaluation Criteria in Solid Tumors (RECIST) from the clinical trial (ctPFS).

**Methods:** Atezolizumab and docetaxel arms of the phase III OAK RCT (NCT02008227) were replicated in a US nationwide real-world database by applying selected OAK inclusion/exclusion criteria, followed by adjustment for baseline prognostic variables using propensity score-based methods. Multiple rwPFS definitions were characterized and a definition was chosen that was acceptable from both clinical and data analysis perspectives. Concordance of outcomes was assessed using Kaplan-Meier (KM) medians and hazard ratios (HRs).

**Results:** Overall, 133 patients receiving atezolizumab and 479 patients receiving docetaxel were selected for the RWD cohort. After adjustment, prognostic variables were balanced between RCT arms and corresponding RWD cohorts. Comparing rwPFS against ctPFS outcomes in terms of KM median and HR showed better concordance for docetaxel (2.99 vs 3.52 months; HR, 0.99, 95% CI, 0.85-1.15) than for atezolizumab (3.71 vs 2.76 months; HR, 0.8, 95% CI 0.61-1.02). The latter improved when events labelled “pseudo-progression” were excluded from the RWD (im-rwPFS) and immune-modified RECIST PFS (im-ctPFS) was used in the RCT Atezolizumab data (4.24 vs 4.14 months; HR, 0.95, 95% CI, 0.70-1.25). These findings were robust across several sensitivity analyses.

**Conclusions:** While rwPFS and ctPFS were similar under docetaxel treatment, this was only the case for atezolizumab when immune-modified progression criteria were used, suggesting that similarity of RWD endpoints to their clinical trial analogues depends on drug category and possibly other factors. Replication of RCTs using RWD and comparison of outcomes can be used as a tool for characterizing RWD endpoints. Additional studies are needed to verify these findings and to better understand the conditions for approximate numerical equivalence of rwPFS and ctPFS endpoints.

## INTRODUCTION

Real-world data (RWD) derived from electronic health records (EHRs) can provide an opportunity to complement clinical trials (CTs) by including broader patient populations that reflect the actual standard of care and by potentially providing access to large amounts of data.[1] These data can help generate insights on all aspects of routine medical care, including the comparison of treatments.

A prerequisite for evaluating therapies using RWD is the availability of reliable and informative endpoints. The analogues of established CT endpoints such as overall survival (OS), progression-free survival (PFS) and overall response rate (ORR) may be extracted from EHRs and used for outcome comparison. However, data collection processes in clinical practice are considerably less standardized and controlled compared to CTs.[2] Therefore, CT endpoints and their respective RWD analogues are not guaranteed to be practically “equivalent”, which implies that they could lead to different numerical results.

Overall survival (OS), the time from randomization to death of any cause, is often considered the “gold standard” endpoint for evaluating cancer therapies.[3] Due to the unambiguous definition of a death event and because close tracking of patients is not required to establish the date of death, OS derived from RWD can be directly compared with OS from CTs, provided the completeness of death information in the RWD database is high.[4-6] This is not generally the case for PFS, a commonly used non-OS endpoint in cancer trials, defined as the time to either progression or death.[7] Data for PFS are collected differently in RWD (rwPFS) compared with CTs (ctPFS), as illustrated in **Figure 1A**. In clinical practice, there are no strict, standardized protocols for scans or scheduled assessments, and patient visits and imaging may occur at irregular time intervals.[8] Patient access to imaging may vary (e.g., due to health insurance coverage). Physicians are also likely to identify clinically relevant progression with the primary objective to make treatment decisions rather than comparing drug efficacy. Hence, it is unclear to what extent real-world progression (rwP) events are captured according to the same criteria as in CTs (Response Evaluation Criteria in Solid Tumors [RECIST]).[8, 9] Continuous patient follow-up, which is required to accurately measure rwPFS, is not always guaranteed in RWD because patients may visit alternative health care providers outside of the data collection network. In addition, there are differences in how mortality data are collected: in CTs, death events are captured up to a clinical cut-off date, which is identical for every patient in the study. Since this does not exist in RWD, a decision must be made on which death events to include in rwPFS. Due to the apparent differences in data collection, a better understanding is needed of when the conditions for comparing rwPFS and ctPFS endpoints in group-level analyses are fulfilled - both in terms of data quality, as well as with respect to indication or treatment category.

**Figure 1.**
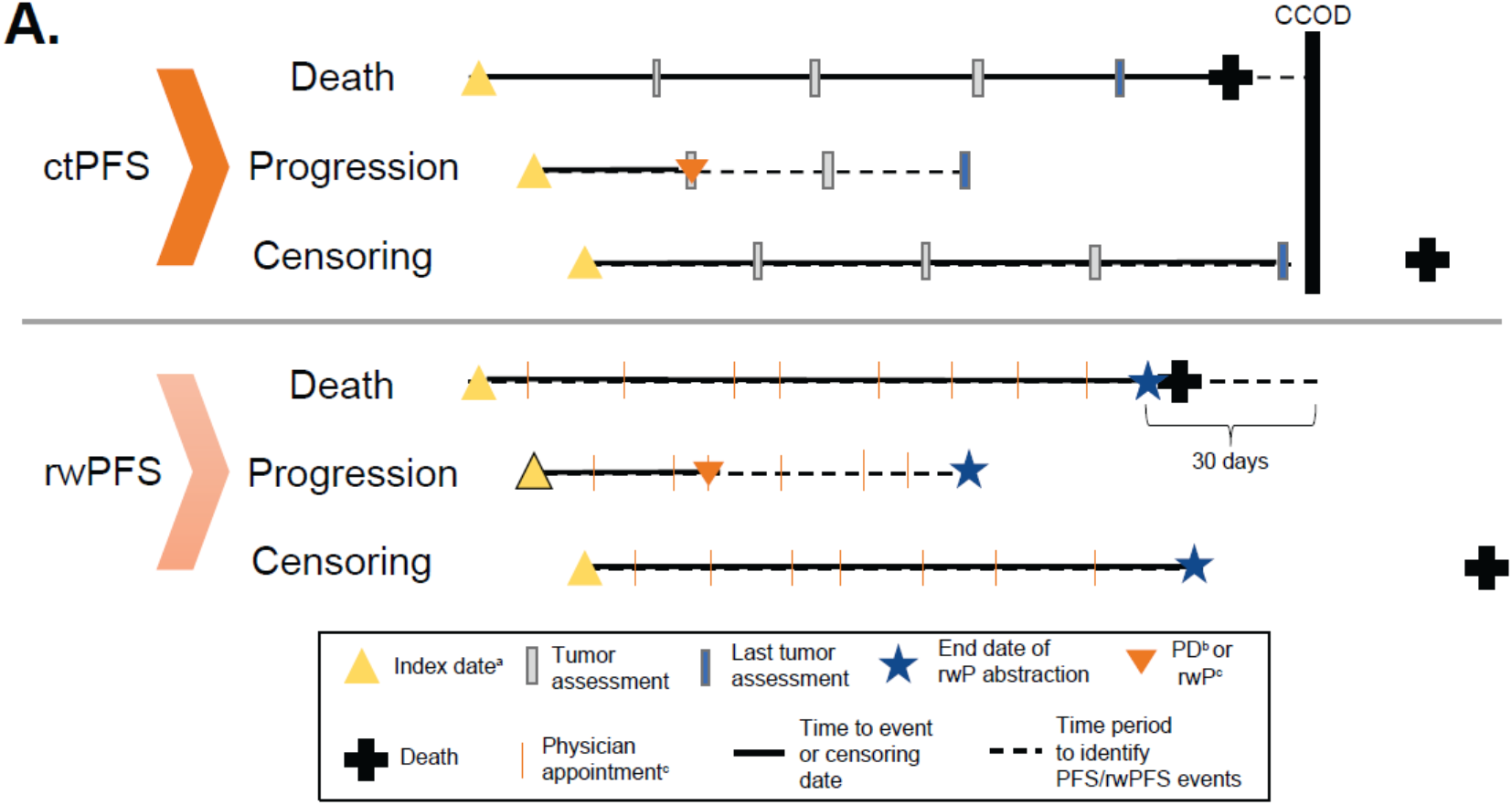
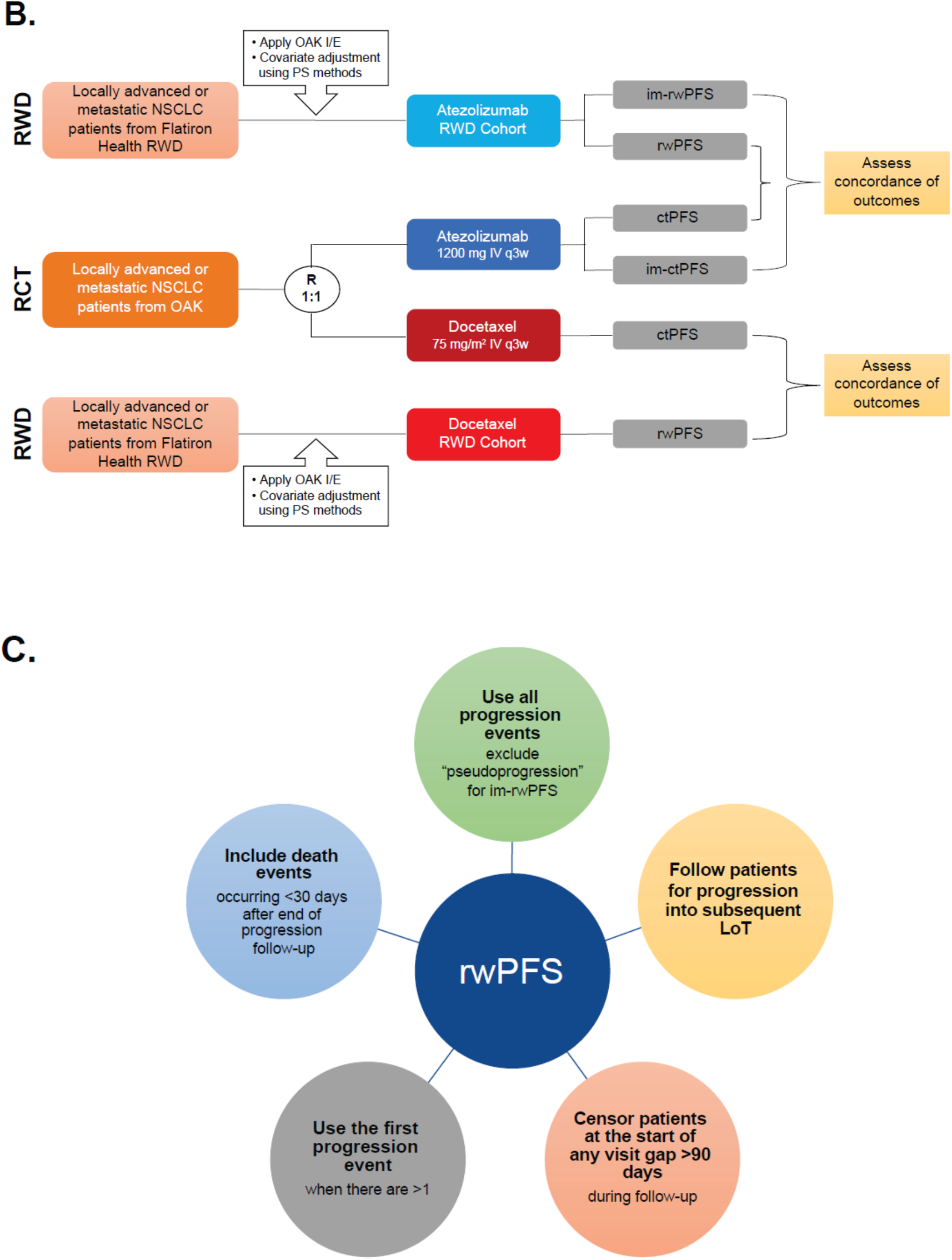
Data collection, study design, and rwPFS definition. A. Data collection processes for CTs and RWD. B. Study design of the OAK RCT and external RWD. C. Proposed definition of rwPFS in this analysis. CCOD, clinical cutoff date; I/E, inclusion/exclusion; im-ctPFS, immune-modified-clinical trial progression-free survival; im-rwPFS, immune-modified-real-world progression-free survival; IV, intravenous; LoT, line of therapy; NSCLC, non-small cell lung cancer; PD, progressive disease; PD-L1, programmed death-ligand 1; PFS, progression-free survival; PS, propensity score; q3w, every 3 weeks; RCT, randomized controlled trial; CT, clinical trial; RWD, real-world data; rwP, real-world progression; rwPFS, real-world progression-free survival. ^a^ Randomization for CT and start of treatment for RWD. ^b^ In CT. ^c^ In RWD.

RWD endpoints, such as time to next treatment, time to treatment discontinuation, time to progression, and rwPFS, have been characterized by assessing surrogacy and correlation of outcomes with OS within the same patients.[10, 11] With this approach it is possible to verify if real-world endpoints are consistent with each other and whether they predict OS. However, such an analysis does not provide any insight into numerical similarity of real-world endpoints and CT analogues. Even an endpoint that perfectly predicts OS could do so by yielding very different numerical results.

Near-equivalence of CT and analogous RWD-derived endpoints is particularly important for comparisons across different datasets, such as in the development of external control (EC) arms from RWD.[12] In this type of analysis, an EC cohort is selected by applying trial inclusion and exclusion criteria to the RWD source, followed by additional covariate adjustment using propensity-score (PS) methods.[13]

Such replication of CT arms from RWD can be used as a tool to assess the comparability of real-world endpoints and their CT analogues. If ctPFS outcomes can reliably and consistently be replicated using rwPFS, this increases confidence in numerical equivalence of the two endpoints - provided good balance of prognostic variables was achieved between the patient populations. A previous study with four CTs for metastatic non-small lung cancer (NSCLC) demonstrated that rwPFS can replicate ctPFS for patients receiving chemotherapy.[14] Another preliminary study in patients with metastatic breast cancer found rwPFS outcomes derived from EHR RWD were similar to ctPFS.[15] The primary aim and novelty of the present analysis was to replicate the cancer immunotherapy (CIT; atezolizumab) treatment arm of the OAK Phase III RCT (using both the ctPFS and immune-modified [im]-ctPFS endpoints of OAK), in addition to the chemotherapy (docetaxel) treatment arm, and to assess concordance of outcomes between cohorts using the rwPFS and ctPFS endpoints.

Finally, there are several different ways of computing rwPFS from the raw data, with little information in the literature describing which methods would most closely replicate ctPFS. Hence, a secondary aim of this analysis was to investigate the impact of varying the rwPFS definition and propose best practices for future, similar analyses.

## METHODS

### Data Source and Study Design

Data from the global, multicenter, open-label, randomized, controlled, Phase III “OAK” trial (NCT02008227),[16, 17] were used to evaluate the ability to replicate ctPFS outcomes in a RWD-derived cohort, using the rwPFS endpoint (**Figure 1B**). Patients with advanced NSCLC (aNSCLC) who had received 1-2 prior lines of chemotherapy (≥1 of which platinum-based) were included, irrespective of programmed death-ligand 1 (PD-L1) expression levels. Patients received either the PD-L1 inhibitor, atezolizumab, 1200 mg intravenously (IV) every 3 weeks (q3w) until progressive disease (PD) or loss of clinical benefit, or the chemotherapeutic drug, docetaxel, 75 mg/m^2^ IV q3w until PD.

ctPFS was determined by the investigator according to RECIST v1.1 guidelines, which comprise pre-specified criteria for defining progression, standard imaging protocols, and scheduled assessments.[9] ctPFS was defined as the time between the date of randomization and the date of first documented PD or death. Participants who were alive and had not experienced PD at the time of analysis were censored at the time of the last tumor assessment. The OAK trial also included im-ctPFS as an exploratory endpoint, which used immune-modified RECIST guidelines (imRECIST) to assess progression. Per imRECIST, a progression is considered a pseudo-progression and not a true progression event if the subsequent scan (≥4 weeks) reads a stable disease, partial response, or complete response.[17, 18]

This study used the US nationwide Flatiron Health (FH) EHR-derived de-identified database to construct external CIT and chemotherapy arms. The FH database is a longitudinal database, comprising de-identified patient-level structured and unstructured data, curated via technology-enabled manual abstraction. During the study period, the de-identified data originated from approximately 280 cancer clinics (≈800 sites of care).[19, 20] Patients included in the database were diagnosed with advanced or metastatic disease on or after January 1, 2011 and had ≥2 visits in the FH system. Inclusion and exclusion criteria from the OAK trial [16] were applied to the data where feasible, and patients who had received atezolizumab or docetaxel and satisfied all criteria were included (**Table S1**). Additionally, patients who did not start first line therapy within 90 days of their advanced diagnosis date were excluded in order to select patients who more closely replicated those enrolled in the RCTs.

### Descriptive statistics and tests

Demographic and clinical characteristics were summarized at the time of index date or the pre-defined baseline period near the index date. Means, standard deviations (SD), medians, interquartile ranges (IQR), and ranges were used to describe continuous variables. Frequencies and proportions were used to describe categorical variables. Variables were compared between the RCT and external CIT arms using Chi-square tests (or Fisher’s exact tests in cases where two-way cell counts were low) for frequencies. For continuous variables, independent t-tests were used for normally distributed variables and Wilcoxon rank-sum tests for variables with a skewed distribution. *P*-values <0.05 were considered statistically significant for both continuous and categorical variables. All statistical analyses were performed using R (v 4.0.0).[21]

### Covariate adjustment and outcome comparisons

Differences between RWD and RCT cohorts in clinical variables (age, sex, race, histology, stage at diagnosis, smoking history, and months since advanced diagnosis) were adjusted using inverse probability of treatment weighting (IPTW). PS for IPTW adjustment were obtained from a logistic regression model estimating one term for every category in the baseline covariates (no interactions) and a cubic spline term for “months since advanced diagnosis.” PD-L1 status was not included in the PS model because the majority of RWD patients did not receive testing (**Figure S1**). For the primary analysis, average treatment effect (ATE) weighting was chosen with removal (trimming) of patients from non-overlapping PS-regions of both RWD and RCT. This minimizes the risk of remaining covariate imbalances after adjustment, thus allowing for a more direct comparison of the endpoints themselves. Average treatment effect for the treated (ATT) weights, with trimming of RWD patients only, were used in one sensitivity analysis to demonstrate the ability to replicate outcomes in patient populations similar to OAK. Differences between baseline characteristics of RCT arm and CIT RWD cohort were assessed by the standardized mean difference (SMD). A SMD <0.1 was an indicator of good covariate balance,[22] and a SMD <0.2 was an indicator of acceptable covariate balance. Concordance of outcomes between RWD and CT endpoints was assessed by hazard ratios (HRs) and Kaplan-Meier (KM) median time to event. Confidence intervals (CIs) were determined by bootstrapping.

The primary analysis included patients with missing Eastern Cooperative Oncology group (ECOG) score but censored patients at the start of any visit gap >90 days to avoid missing rwP events due to interrupted follow-up. Sensitivity analyses were performed 1) using a doubly robust approach (estimation of a conditional HR by fitting multivariable Cox regression to the IPTW adjusted dataset using the same covariates as the PS model); 2) adjusting for ECOG (excluding patients with missing ECOG); 3) omitting patients who were lost to follow-up <3 months after index date; and 4) using a 60-day time window to capture death events after the end of rwP follow-up (instead of 30 days). Exploratory analyses included 1) estimating the ATT (without IPTW weighting and trimming of RCT patients); and 2) allowing for visit gaps of any length during rwP follow-up (not censoring at the start of 90-day visit gaps).

### Development of an rwPFS Endpoint

While the high-level definition of PFS as time to death or progression is straightforward, several definitions are possible when considering how to compute rwPFS from RWD (**Supplemental Methods**). The impact of the exact rwPFS definition on outcomes was evaluated through tabulation of KM medians, HRs, and event frequencies. Based on the analyses performed, a “reference” definition of rwPFS was chosen that was acceptable from both clinical and data analysis points of view and minimized potential biases (**Table 1, Figure 1C**).

**Table 1.** Elements of ctPFS and corresponding adaptations for RWD. CCOD, clinical cutoff date; EHR, electronic health record; FH, Flatiron Health; FU, follow-up; PD, progressive disease; RCT, randomized controlled trial; RWD, real-world data; rwP, real-world progression. ^a^ Sensitivity analysis was conducted using rwP from radiographic evidence only.

| Criteria | OAK RCT | Adaptation in RWD | Rationale for adaptation |
| --- | --- | --- | --- |
| Time of censoring in absence of an event | Time of last tumor assessment | End of gapless rwP FU.<br>Earliest of:<br>- last recorded visit<br>- start of a visit gap >90 days<br>- last date of rwP abstraction | ● To avoid missing rwP events, it is necessary to ensure patients have continuous FU and rwP is abstracted from the EHR data during the entire FU period. |
| PD event | First PD event by radiographic evaluation | First rwP event (any) during rwP FU | ● Most progression events are radiographically confirmed<br>● Risk of missing rwP events if restricting to radiographically confirmed rwP events only<sup>a</sup> |
| Death event | Death between randomization and CCOD | Death between index date and 30 days after end of rwP FU | ● Patients often leave EHR network shortly before death; hence, death must be captured during a short period after last visit<br>● Pragmatic choice of 30 days captures a large proportion of excess deaths shortly after end of progression FU while still being short (to minimize bias) |
| Absence of event | Alive and have not had PD between randomization and CCOD | Alive at end of rwP FU (+30 days) and have not had rwP |  |

## RESULTS

Overall, 133 patients who received atezolizumab and 479 patients who received docetaxel were selected for the RWD cohorts. Following adjustments via IPTW, 134.7 and 475.1 patients were included in the RWD cohorts for atezolizumab and docetaxel, respectively. Fractional patient number occurred in the adjusted cohorts due to reweighting of data points (**Table 2**).

**Table 2.** Baseline characteristics of the atezolizumab and docetaxel RCT arms and respective RWD cohorts before and after covariate adjustment via inverse probability weighting and trimming. RCT, randomized controlled trial; RWD, real-world data; SMD, standardized mean difference. ^a^ Fractional patient numbers may occur after propensity score adjustments due to reweighting of data points.

| Treatment Arm |  | Atezolizumab |  |  |  |  |  | Docetaxel |  |  |  |  |  |
| --- | --- | --- | --- | --- | --- | --- | --- | --- | --- | --- | --- | --- | --- |
| Variable, n (%) | Categories | Unadjusted |  | SMD | Adjusted |  | SMD | Unadjusted |  | SMD | Adjusted |  | SMD |
|  |  | RCT n=425 | RWD n=133 |  | RCT n=388.4<sup>a</sup> | RWD n=134.7<sup>a</sup> |  | RCT n=425 | RWD n=479 |  | RCT n=413.8<sup>a</sup> | RWD n=475.1<sup>a</sup> |  |
| Age group | <65 | 248.0 (58.4) | 49.0 (36.8) | 0.48 | 201.7 (51.9) | 67.6 (50.2) | 0.05 | 236.0 (55.5) | 199.0 (41.5) | 0.35 | 202.3 (48.9) | 231.5 (48.7) | 0.01 |
|  | 65-74 | 137.0 (32.2) | 50.0 (37.6) |  | 136.4 (35.1) | 50.3 (37.3) |  | 159.0 (37.4) | 195.0 (40.7) |  | 162.7 (39.3) | 186.4 (39.2) |  |
|  | ≥75 | 40.0 (9.4) | 34.0 (25.6) |  | 50.2 (12.9) | 16.9 (12.5) |  | 30.0 (7.1) | 85.0 (17.7) |  | 48.8 (11.8) | 57.2 (12.0) |  |
| Sex | Female | 164.0 (38.6) | 64.0 (48.1) | 0.18 | 158.8 (40.9) | 51.8 (38.4) | 0.05 | 166.0 (39.1) | 212.0 (44.3) | 0.09 | 173.2 (41.8) | 194.0 (40.8) | 0.02 |
|  | Male | 261.0 (61.4) | 69.0 (51.9) |  | 229.6 (59.1) | 83.0 (61.6) |  | 259.0 (60.9) | 267.0 (55.7) |  | 240.7 (58.2) | 281.1 (59.2) |  |
| Histology | Non-squamous | 313.0 (73.6) | 101.0 (75.9) | 0.03 | 294.0 (75.7) | 108.9 (80.9) | 0.12 | 315.0 (74.1) | 360.0 (75.2) | 0.03 | 307.8 (74.4) | 356.1 (75.0) | 0.01 |
|  | Squamous | 112.0 (26.4) | 32.0 (24.1) |  | 94.3 (24.3) | 25.8 (19.1) |  | 110.0 (25.9) | 119.0 (24.8) |  | 106.0 (25.6) | 119.0 (25.0) |  |
| Stage at diagnosis | I | 24.0 (5.6) | 16.0 (12.0) | 0.35 | 28.0 (7.2) | 9.0 (6.6) | 0.03 | 34.0 (8.0) | 31.0 (6.5) | 0.23 | 30.1 (7.3) | 33.9 (7.1) | 0.01 |
|  | II | 31.0 (7.3) | 5.0 (3.8) |  | 23.3 (6.0) | 8.6 (6.4) |  | 23.0 (5.4) | 22.0 (4.6) |  | 19.5 (4.7) | 22.2 (4.7) |  |
|  | IIIA | 61.0 (14.4) | 5.0 (3.8) |  | 34.2 (8.8) | 11.2 (8.3) |  | 52.0 (12.2) | 30.0 (6.3) |  | 36.9 (8.9) | 42.7 (9.0) |  |
|  | IIIB | 53.0 (12.5) | 20.0 (15.0) |  | 54.2 (14.0) | 19.1 (14.2) |  | 55.0 (12.9) | 53.0 (11.1) |  | 46.7 (11.3) | 54.3 (11.4) |  |
|  | IV | 247.0 (58.1) | 87.0 (65.4) |  | 248.7 (64.0) | 86.9 (64.5) |  | 257.0 (60.5) | 337.0 (70.4) |  | 276.4 (66.8) | 317.3 (66.8) |  |
|  | Unknown | 9.0 (2.1) | 0.0 (0.0) |  | 0 | 0 |  | 4.0 (0.9) | 6.0 (1.3) |  | 4.3 (1.0) | 4.8 (1.0) |  |
| Smoking history | History | 341.0<br>(80.2) | 114.0<br>(85.7) | 0.13 | 317.9<br>(81.9) | 107.2<br>(79.6) | 0.06 | 353.0<br>(83.1) | 434.0<br>(90.6) | 0.20 | 361.5<br>(87.3) | 409.1<br>(86.1) | 0.04 |
|  | No history | 84.0<br>(19.8) | 19.0<br>(14.3) |  | 70.5<br>(18.1) | 27.5<br>(20.4) |  | 72.0<br>(16.9) | 45.0<br>(9.4) |  | 52.3<br>(12.7) | 66.0<br>(13.9) |  |
| Race | White | 302.0<br>(71.1) | 101.0<br>(75.9) | 0.17 | 282.8<br>(72.8) | 101.3<br>(75.2) | 0.05 | 296.0<br>(69.6) | 334.0<br>(69.7) | 0.16 | 291.8<br>(70.5) | 329.6<br>(69.4) | 0.03 |
|  | Others | 103.0<br>(24.2) | 22.0<br>(16.5) |  | 84.3<br>(21.7) | 26.6<br>(19.8) |  | 115.0<br>(27.1) | 110.0<br>(23.0) |  | 102.1<br>(24.7) | 123.1<br>(25.9) |  |
|  | Unknown | 20.0<br>(4.7) | 10.0<br>(7.5) |  | 21.3<br>(5.5) | 6.8 (5.1) |  | 14.0<br>(3.3) | 35.0<br>(7.3) |  | 19.9<br>(4.8) | 22.5<br>(4.7) |  |
| Months since advanced diagnosis, mean (SD) |  | 15.47<br>(14.49) | 12.26<br>(10.31) | 0.17 | 13.82<br>(12.68) | 14.87<br>(13.48) | 0.08 | 13.90<br>(13.47) | 9.53<br>(5.79) | 0.37 | 11.01<br>(7.79) | 11.27<br>(8.17) | 0.03 |

### Replication of the OAK Atezolizumab Arm

Good balance of prognostic variables was achieved between the atezolizumab RWD cohort and the corresponding RCT arm. After IPTW ATE weighting and trimming, SMD values were <0.1 with the exception of histology (0.12) (**Table 2**). KM median time-to-event was longer in rwPFS compared with ctPFS (rwPFS: 3.71, 95% CI, 2.5-4.83 months vs ctPFS: 2.76 95% CI, 2.43-2.96 months; **Figure 2A**). The corresponding HR indicated a lower event rate for rwPFS (HR, 0.80, 95% CI, 0.61-1.02). A similar pattern was observed consistently across all sensitivity analyses (**Figure 2B**). Among patients who had a PFS event, 12.7% were death events in rwPFS, compared with 13.1% in ctPFS. When events labelled “pseudo-progression” were omitted in the RWD (im-rwPFS) and compared with RCT PFS evaluated according to imRECIST guidelines (im-ctPFS), the KM median time-to-event was similar (im-rwPFS: 4.24, 95% CI, 2.83-6.64 months vs im-ctPFS: 4.14, 95% CI, 3.48-4.63 months) and the corresponding HR was close to unity (HR, 0.95, 95% CI, 0.70-1.25; **Figure 2C**). A similar pattern was observed consistently across all sensitivity analyses (**Figure 2D**). The rwPFS and im-rwPFS endpoints assessed by KM curves were very similar **(Figure S2)**. Among patients who had a PFS event, 14.0% were death events in im-rwPFS, compared with 25.8% in im-ctPFS.

**Figure 2.**
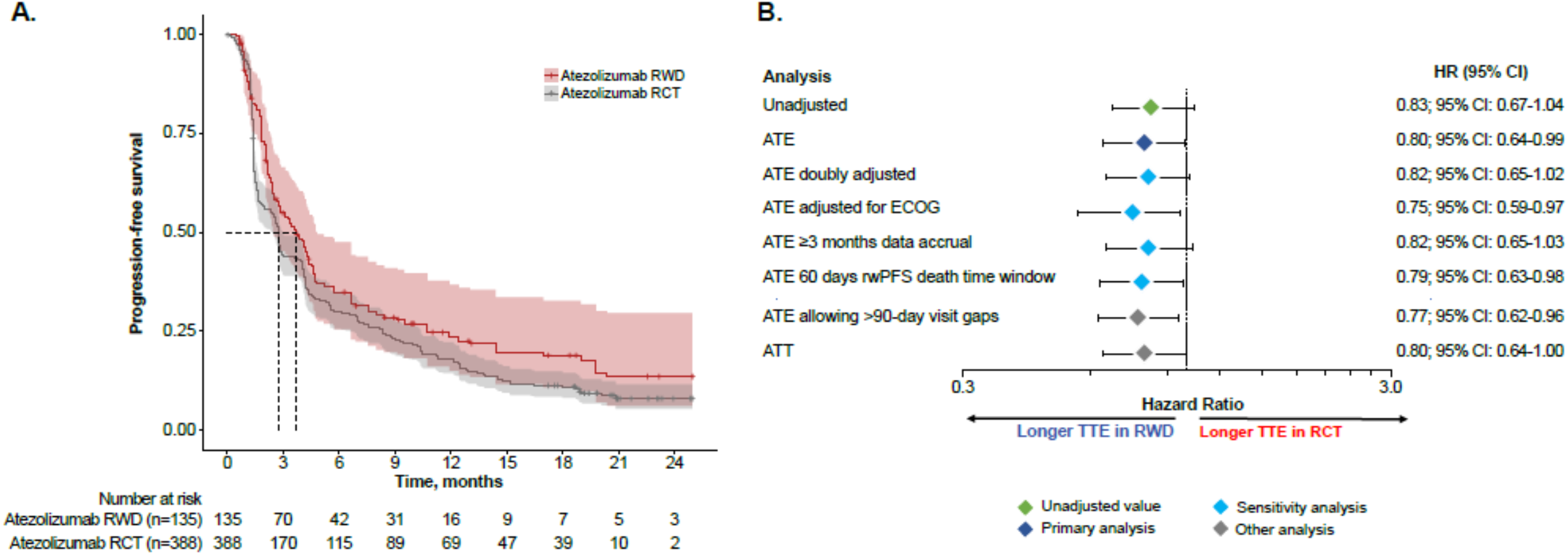
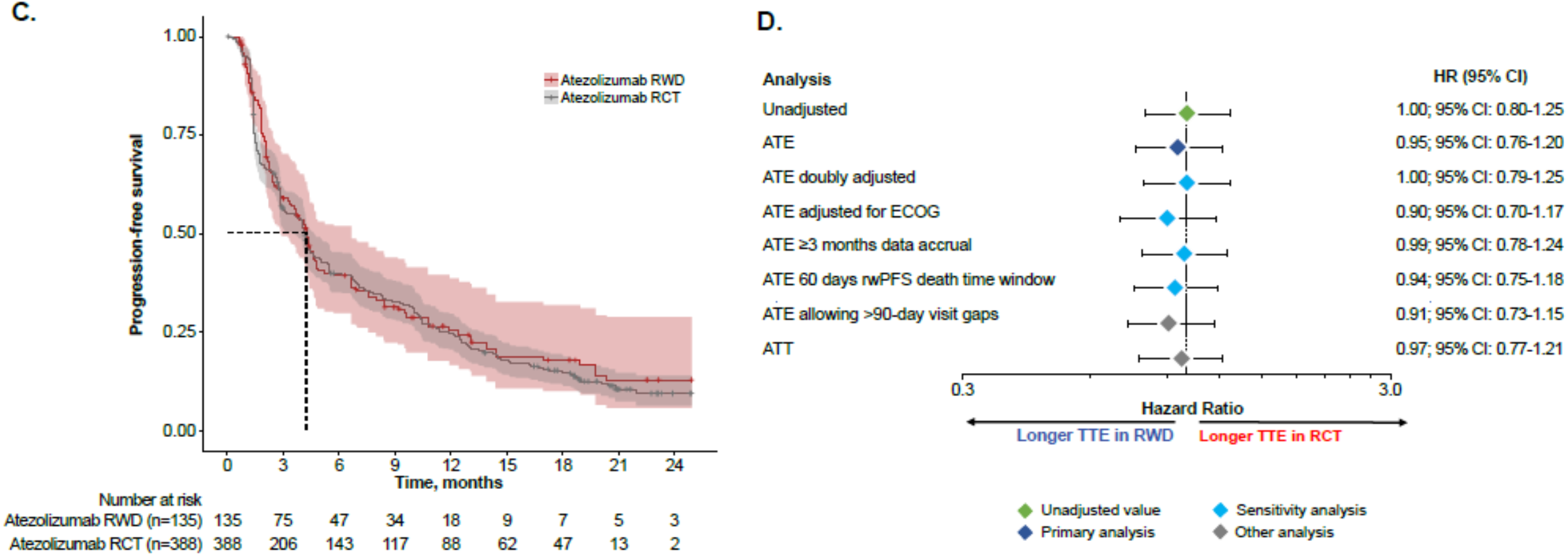
Replication of the OAK RCT atezolizumab arm. A. KM curves of ctPFS and rwPFS and B. corresponding HR values. C. KM curves of im-ctPFS and im-rwPFS and D. corresponding HR values. ATE, average treatment effect; ATT, average treatment effect in the treated; ECOG, Eastern Cooperative Oncology Group performance score; HR, hazard ratio; PFS, progression-free survival; im-ctPFS, immune-modified clinical trial progression-free survival; im-rwPFS, immune-modified real-world progression-free survival; RCT, randomized controlled trial; ctPFS, clinical trial progression-free survival; rwPFS, real-world progression-free survival; TTE, time to event

### Replication of the OAK Docetaxel Arm

Good balance of prognostic variables was achieved between the docetaxel RWD cohort and the corresponding RCT arm. After IPTW ATE weighting and trimming all SMD values were <0.1 (**Table 2**). KM median time-to-event was similar in rwPFS compared with ctPFS (rwPFS: 2.99, 95% CI, 2.56-3.58 months vs ctPFS: 3.52 95% CI, 2.89-4.14 months; **Figure 3A**). The corresponding HR was close to unity (HR, 0.99, 95% CI, 0.85-1.15), with a similar pattern observed consistently across all sensitivity analyses (**Figure 3B**). Among patients who had a PFS event, 16.6% were death events in rwPFS, compared with 23.5% in ctPFS.

**Figure 3.**
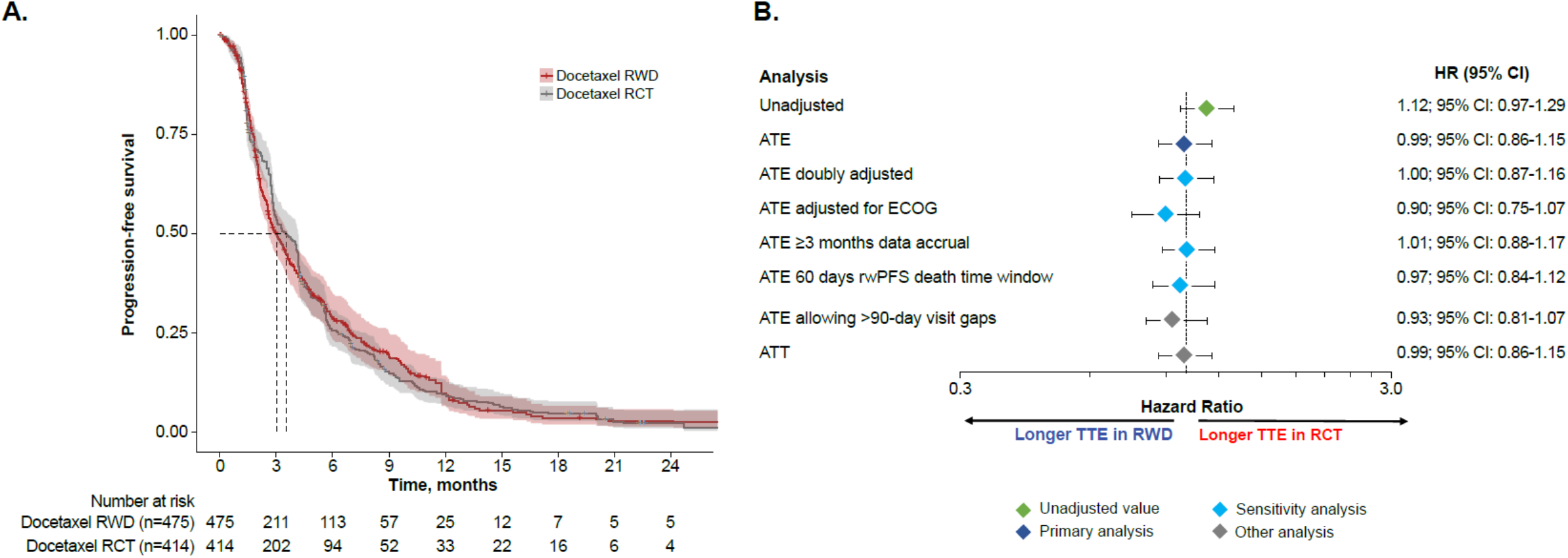
Replication of the OAK docetaxel arm. A. KM curves of RWD rwPFS and OAK PFS. B. HRs by analysis. ATE, average treatment effect; ATT, average treatment treated; ECOG, Eastern Cooperative Oncology Group; HR, hazard ratio; KM, Kaplan Meier; PFS, progression-free survival; RCT, randomized controlled trial; RECIST, response evaluation criteria in solid tumors; rwPFS, real-world progression-free survival; TTE, time to event.

### Characterization of rwPFS

Follow-up of RWD patients was close, with clinic visits occurring frequently. Gaps longer than 2-3 weeks between two visits or between last visit and rwPFS event/censoring were rare (**Figure S3**). Altering the rwPFS definition by using different subsets of rwP events and by varying the handling of line of therapy (LoT) change (**Table 3**) resulted overall in only minor outcome changes compared to the “reference” endpoint definition. The largest change in the atezolizumab cohort was observed when excluding events labelled “pseudo-progression” (KM median 4.1 months vs 3.58 months; HR 0.93; **Figure S2**). In the docetaxel cohort, the largest change was observed when imputing a progression event at the start of a new LoT (KM median 2.53 months vs 2.76 months; HR 1.17). The time window for capturing death events after the end of progression follow-up was varied from 0-60 days, in 10-day increments, and the impact on event composition, KM median time-to-event, and HR (relative to the “reference” definition) was recorded (**Table S2**). In the docetaxel cohort, the number of additional death events captured per 10-day increment was largest for window sizes up to 30-40 days and decreased thereafter. A similar pattern was observed for KM median and HR, which showed marked sensitivity to window size at small absolute values, with changes per increment decreasing at window sizes above 30-40 days. The number of death events in the atezolizumab cohort was too small to draw firm conclusions, but results appeared consistent with the pattern observed in the docetaxel cohort.

**Table 3.**
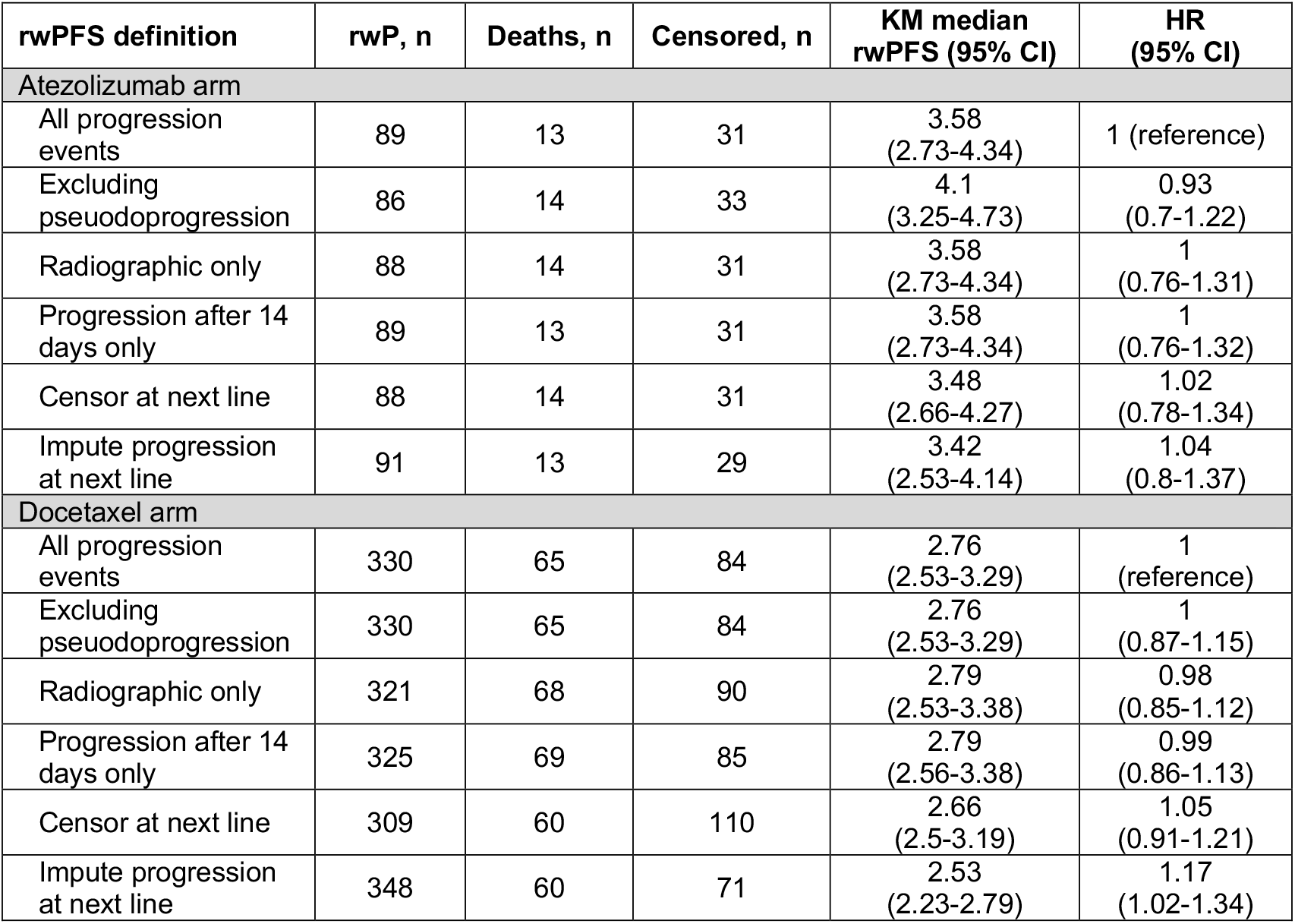
Effects of varying the event and censoring definitions. CI, confidence interval; HR, hazard ratio; KM, Kaplan Meier; rwP, real-world progression; rwPFS, real-world progression-free survival.

### DISCUSSION/CONCLUSION

This study investigated the concordance of PFS outcomes in the OAK trial with those from matched real-world cohorts in patients diagnosed with aNSCLC. A novel aspect and focus of this analysis was the assessment in patients receiving a CIT regimen, since many future experimental drugs will likely need to be compared against a CIT standard of care. The main objective was to better understand the extent to which clinical trial PFS and its real-world analogue(s) could be considered equivalent endpoints, meaning that they would lead to the same or very similar numerical results in any group-level statistical comparison. EHR-derived cohorts were selected for both the atezolizumab and docetaxel arms of OAK. A good balance of prognostic variables was achieved after IPTW ATE and trimming (removing) patients from both trial data and RWD. This is necessary for interpreting concordance of outcomes as potentially indicative of a high similarity between real-world and trial endpoints.

Very high concordance was observed between rwPFS and ctPFS outcomes in docetaxel-treated patients (**Figure 3**). This is consistent with the PFS results of a similar study in aNSCLC where the chemotherapy control arms of four clinical trials were replicated using RWD.[14] Together, these results increase the confidence that rwPFS and ctPFS endpoints are largely equivalent in aNSCLC patients under a chemotherapy regimen. A different observation was made for patients receiving atezolizumab: median rwPFS was approximately one month longer compared to median ctPFS. Interestingly, very good concordance was observed in the same patients when im-rwPFS and im-ctPFS endpoints were compared, using immune-modified progression criteria in both RWD and RCT data. This change can largely be attributed to the difference between im-ctPFS vs ctPFS because the difference between im-rwPFS and rwPFS appeared small in comparison (**Figure 2 A-B, Figure S2, Table 3**). These observations raise the possibility that rwPFS under CIT treatment may correspond more closely to im-RECIST PFS, but it remains unclear whether this is a general pattern or an observation unique to this analysis. If a general pattern, this would imply that caution should be exercised when using rwPFS to compare RWD of CIT treatments with CT data, or to compare a CIT against other drug categories within RWD. However, if the present RWD Atezolizumab cohort were used as external control arm for a single arm clinical trial, the direction of observed bias (longer time to event) suggests a conservative estimate of PFS benefit for the experimental treatment. Importantly, these findings indicate that the similarity of rwPFS and ctPFS endpoints may be drug dependent. Therefore, concordance of outcomes may need to be assessed separately for different drug categories. Similarly, concordance of rwPFS and ctPFS outcomes should also be verified empirically in other cancers.

Ideally, confidence in real-world endpoints could be fully established via a bottom-up approach, similar to clinical trials, where a thorough understanding of data generation provides reassurance of outcome validity. Due to the complex and poorly understood processes that generate RWD, this is likely not possible, and empirical evidence similar to the present study will likely continue to play an important role. Nevertheless, attempts should be made to characterize and understand data generation in RWD as well as possible, through detailed analysis of tumor imaging data, temporal patterns of clinic visits, or qualitative research where physicians describe what led to the decisions recorded in the data. Such analyses may help understand whether apparent equivalence of endpoints on a group level implies equivalence at the individual level. Despite differences in data collection between CT and RWD, it is possible that rwPFS and ctPFS may yield similar results. This could be the result of a shared primary objective, where both processes in routine health care and in clinical trials were designed to identify clinically relevant progression events. Different secondary objectives may be present, such as physicians making treatment decisions rather than comparing effectiveness, or clinical trials being designed to be auditable. Additionally, minimizing cost and reducing unnecessary discomfort for patients through frequent imaging may be other factors considered in routine clinical care.

During the planning of this study, it became apparent that there are multiple ways of computing rwPFS from RWD, and no consensus could be reached on a single “correct” rwPFS definition (**Supplemental information**). Instead, a pragmatic choice from a range of acceptable definitions was made, and the implications of varying the definition on rwPFS outcomes were documented (**Table 3, Table S2**) in line with FDA draft guidelines.[23] The endpoint definition did not affect outcomes strongly enough to substantially alter the study results, with the following exceptions: 1) the omission of events labelled “pseudo-progression” for patients receiving atezolizumab as discussed above, 2) the imputation of a progression event at LoT change for patients receiving docetaxel, and 3) the size of the time window for capturing death events after the end of progression follow-up. From a clinician’s point of view, a new LoT is initiated after the patient has progressed, and consequently imputing a progression event at LoT change should not have made a substantial difference. It is unclear why there was an effect among the docetaxel patients, but it may suggest an inconsistency between the LoT and progression information in the database. The decision to choose a 30-day time window for capturing death events was motivated by the observation that extending the window size from 0 to 30 days had a substantial effect on HRs (in line with a hypothesis of patients leaving the FH network shortly before death), while extending the window size further resulted in minimal change. Software in the form of an R package (supplemental methods) is provided that simplifies calculation of rwPFS, as well as the comparison of different definitions.[24]

Some limitations of this study are worth noting. Because the study was restricted to patients with aNSCLC who were mostly from community settings, results may not be generalizable to other tumor types, stages, and patient populations. PD-L1 status was not used for PS weighting due to a limited number of real-world patients who received any PD-L1 test. Patients in both RWD atezolizumab and docetaxel treatment groups did not start their treatment during the same period in which trial patients were recruited, which has the potential to introduce calendar time bias due to changes in the standard of care. Any conclusions regarding rwPFS in atezolizumab should be considered preliminary as they are based on the replication of a single trial arm. Common limitations of RWD studies apply, where unmeasured confounding factors and missing data can still exist [13].

In conclusion, ctPFS outcomes could successfully be replicated using rwPFS for docetaxel, while replication was better for atezolizumab when immune-modified progression criteria were used in both RWD and RCT data. Additional studies are needed to verify these findings across different CITs approved for aNSCLC and in other indications.

## Data Availability

Qualified researchers may request access to individual patient level data through the
clinical study data request platform (https://vivli.org/). Further details on Roche's criteria for
eligible studies are available here (https://vivli.org/members/ourmembers/). For further details on
Roche's Global Policy on the Sharing of Clinical Information and how to request access to
related clinical study documents, see here
(https://www.roche.com/research_and_development/who_we_are_how_we_work/clinical_trials/our_commitment_to_data_sharing.htm).
The data that support the findings of this study originated from Flatiron Health, Inc.
These de-identified data may be made available upon request and are subject to a license
agreement with Flatiron Health; interested researchers should contact
to determine licensing terms.

https://github.com/phcanalytics/RwPFS

## ACKNOWLEDGMENTS

We thank the patients who participated in this study and their families. This manuscript was sponsored by F. Hoffmann-La Roche Ltd. and Genentech, Inc. Support for third-party writing assistance, furnished by Jessica Swanner, PhD, of Health Interactions, Inc., was provided by F. Hoffmann-La Roche Ltd. We thank many of our colleagues for their helpful discussions surrounding this work.

## DATA SHARING STATEMENT

Qualified researchers may request access to individual patient level data through the clinical study data request platform (https://vivli.org/). Further details on Roche’s criteria for eligible studies are available here (https://vivli.org/members/ourmembers/). For further details on

Roche’s Global Policy on the Sharing of Clinical Information and how to request access to related clinical study documents, see here (https://www.roche.com/research_and_development/who_we_are_how_we_work/clinical_trials/our_commitment_to_data_sharing.htm).

The data that support the findings of this study originated from Flatiron Health, Inc. These de-identified data may be made available upon request and are subject to a license agreement with Flatiron Health; interested researchers should contact to determine licensing terms.

## AUTHOR CONTRIBUTION STATEMENTS

All authors had full access to the data in the study and take responsibility for the integrity and the accuracy of the data analysis. All authors contributed substantially to the study design (SKM, TGNT, MTB, RJMM, HT), data analysis (MTB, RJMM, HT), interpretation of results (SKM, MTB, TGNT, HT, JM, AR), and the development of the manuscript (MTB, SKM, TGNT, HT, JM, AR). All authors read and approved the final manuscript.

## SUPPLEMENTAL INFORMATION

### Methods

The following considerations illustrate the range of possible real-world progression-free survival (rwPFS) definitions:

- <u>rwP events</u>: i) Should all types of real-world progression (rwP) events in the Flatiron Health (FH) database be used?, or ii) should events flagged as “pseudo-progression” be omitted?, or iii) should only radiographically confirmed events be used (in line with the clinical trial data)?, and should v) events happening within 14 days of index date be discarded (as per FH recommendation)?
- <u>Line of Therapy (LoT) change</u>: From a clinician’s point-of-view, a new LoT is associated with progression. However, the reason for LoT change is usually not available in real-world data (RWD). Questions: i) should patients be followed for rwP beyond LoT change?, or ii) should patients be censored at LoT change?, or iii) should a progression event be imputed at LoT change (in line with the clinical LoT concept)?
- <u>Continuous follow-up</u>: Unlike overall survival (OS), progression-free survival (PFS) requires continuous follow-up. In RWD this may not always be the case, as patients may leave the FH network and seek treatment elsewhere. Hence, it may be beneficial to censor patients at the start of long visit gaps (indicating absence) to avoid missing rwP events. Question: how long should the maximally permissible gap in recorded visits be?
- <u>Death events</u>: Typically, patients leave the FH network shortly before death (i.e., transferred to hospice or a hospital). This requires capturing death events for some period after end of progression follow-up. How large should the time window be to avoid informative censoring? A larger time window captures more death events but is also more prone to the following two biases: i) missing rwP events, leading to underestimation of the event rate, and ii) differential treatment (censoring at end of rwP follow-up vs recording of a death event) of patients depending on experiencing a death event or not.^19^

**<u>RwPFS R package</u>** to calculate and compare a range of rwPFS endpoints with varying definitions.

**Table S1.**
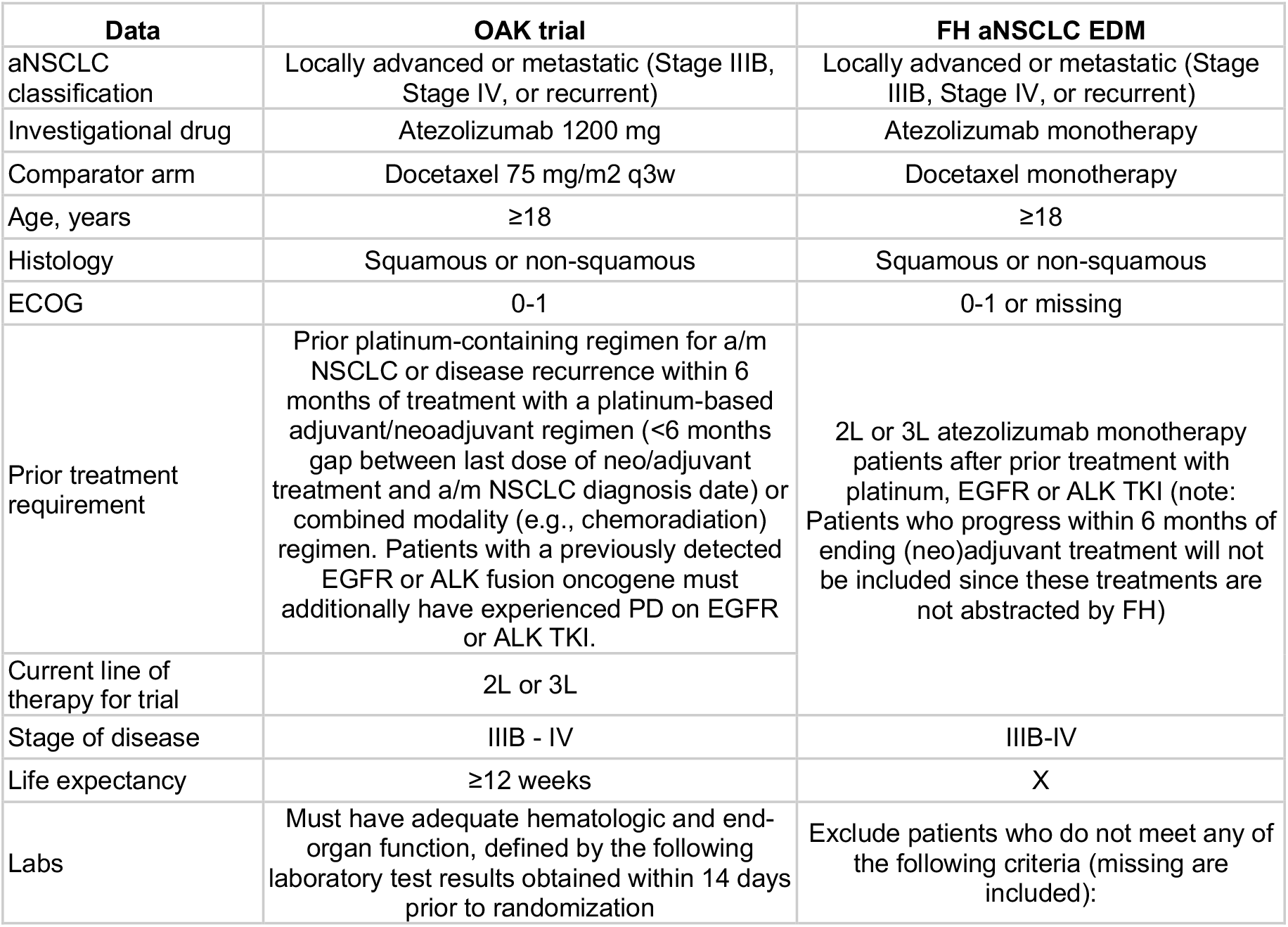
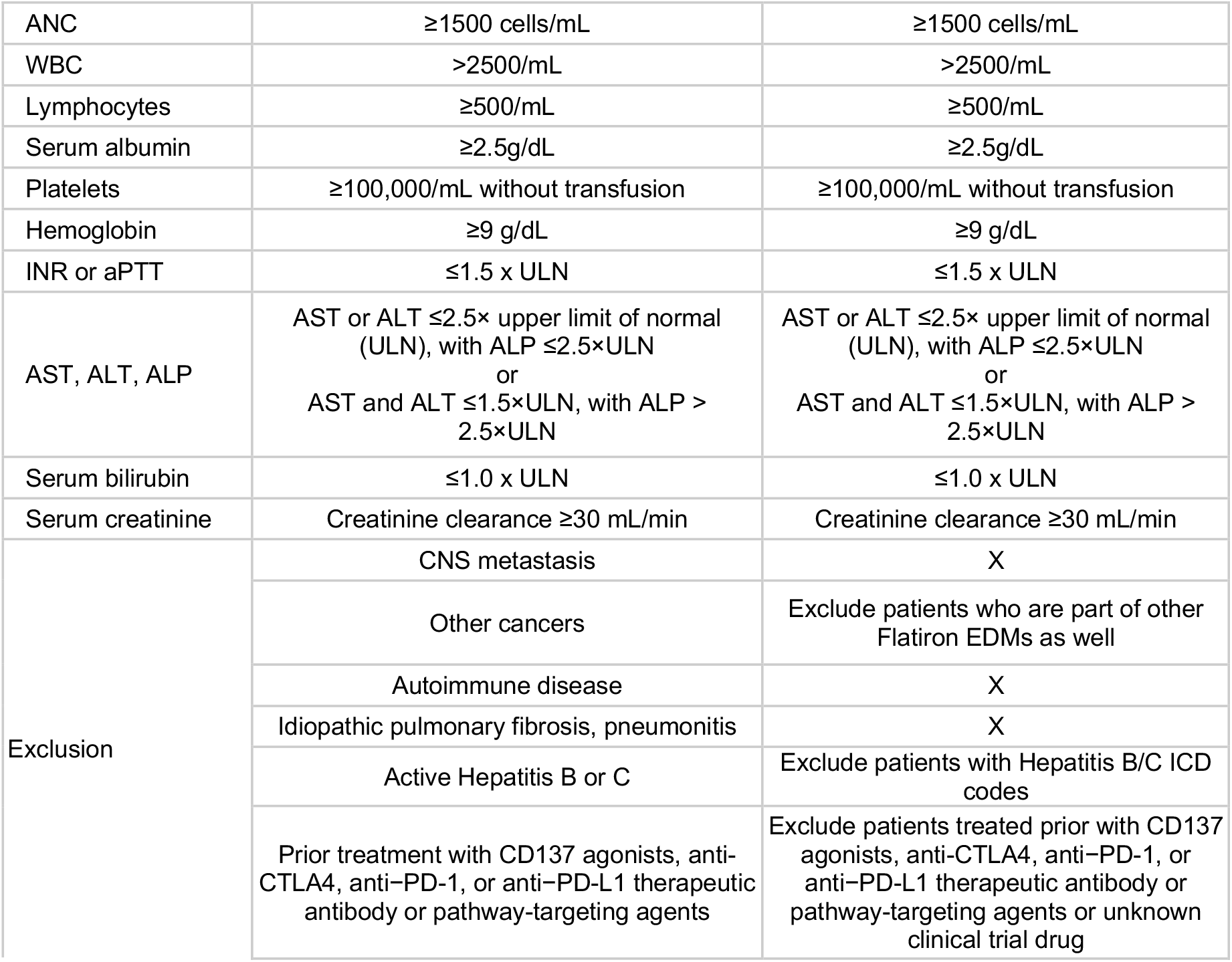
Inclusion/Exclusion Criteria From the OAK Trial Applied to the FH aNSCLC EDM. 2L, second-line; 3L, third-line; ALK, anaplastic lymphoma kinase; ALP, alkaline phosphatase; a/m, advanced/metastatic; aNSCLC, advanced non-small cell lung cancer; ALT, alanine aminotransferase; ANC, absolute neutrophil count; aPPT, activated partial thromboplastin time; AST, aspartate aminotransferase; CD137, tumor necrosis factor receptor superfamily, member 9; CNS, central nervous system; CTLA4, Cytotoxic T lymphocyte-associated antigen-4; EDM, enhanced data mart; EGFR, epidermal growth factor receptor; FH, Flatiron Health; ICD, international classification of disease; INR, international normalized ratio; PD-1, programmed death 1; PD-L1, programmed death-ligand 1; q3w, every 3 weeks; TKI, tyrosine kinase inhibitor; UNL, upper limit of normal; WBC, white blood cell count; X, Application of the criteria to RWD was not feasible.

**Table S2.** Effects of varying the time window for capturing death after end of rwP follow-up. CI, confidence interval; HR, hazard ratio; KM, Kaplan Meier; rwP, real-world progression; rwPFS, real-world progression-free survival.

| Death time window, days | rwP, n | Deaths, n (%) | Censored, n | Additional deaths, n | KM median rwPFS (95% CI) | HR (95% CI) |
| --- | --- | --- | --- | --- | --- | --- |
| **Atezolizumab arm** |  |  |  |  |  |  |
| 0 | 89 | 1 (1.1) | 43 |  | 4.07<br>(3.35-4.83) | 0.89<br>(0.67-1.19) |
| 10 | 89 | 5 (5.3) | 39 | 4 | 3.81<br>(3.25-4.67) | 0.93<br>(0.7-1.23) |
| 20 | 89 | 8 (8.2) | 36 | 3 | 3.68<br>(2.89-4.63) | 0.96<br>(0.73-1.27) |
| 30 | 89 | 13 (12.7) | 31 | 5 | 3.58<br>(2.73-4.34) | 1<br>(reference) |
| 40 | 89 | 15 (14.4) | 29 | 2 | 3.42<br>(2.66-4.27) | 1.01<br>(0.77-1.33) |
| 50 | 89 | 17 (16.0) | 27 | 2 | 3.42<br>(2.66-4.27) | 1.03<br>(0.78-1.35) |
| 60 | 89 | 18 (16.8) | 26 | 1 | 3.35<br>(2.53-4.24) | 1.03<br>(0.79-1.36) |
| **Docetaxel arm** |  |  |  |  |  |  |
| 0 | 330 | 9 (2.7) | 140 |  | 3.35<br>(2.79-3.98) | 0.87<br>(0.76-1.01) |
| 10 | 330 | 25 (7.0) | 124 | 16 | 3.09<br>(2.69-3.71) | 0.91<br>(0.79-1.05) |
| 20 | 330 | 44 (11.8) | 105 | 19 | 2.89<br>(2.56-3.45) | 0.96<br>(0.83-1.1) |
| 30 | 330 | 65 (16.5) | 84 | 21 | 2.76<br>(2.53-3.29) | 1<br>(reference) |
| 40 | 330 | 78 (19.1) | 71 | 13 | 2.66<br>(2.5-3.19) | 1.02<br>(0.89-1.17) |
| 50 | 330 | 84 (20.3) | 65 | 6 | 2.66<br>(2.5-3.09) | 1.03<br>(0.9-1.18) |
| 60 | 330 | 93 (22.0) | 56 | 9 | 2.66<br>(2.5-3.09) | 1.04<br>(0.91-1.2) |

**Figure S1.**
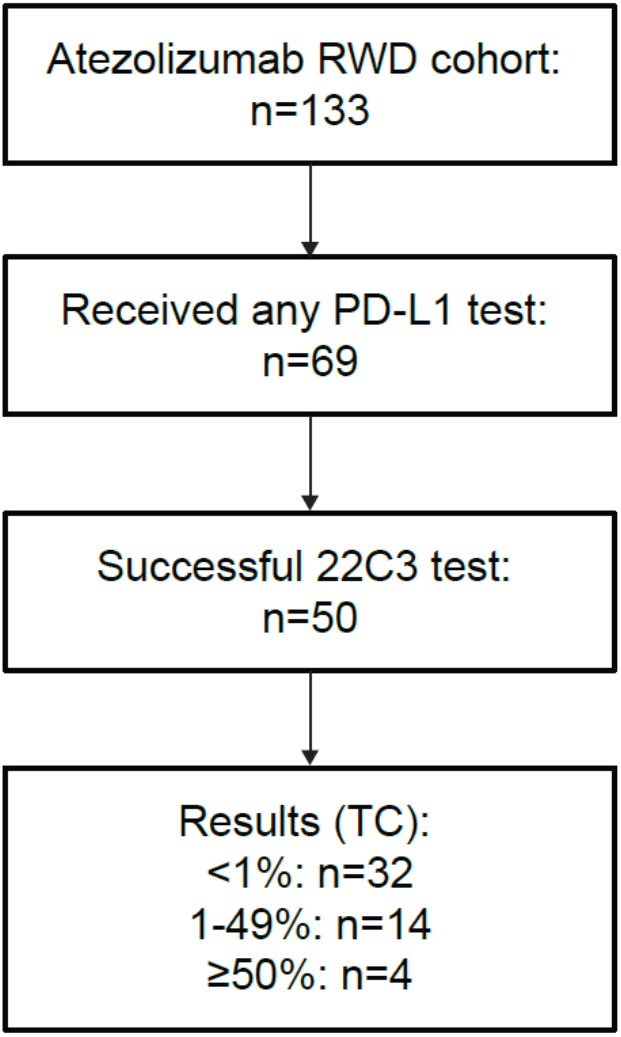
PD-L1 testing in the atezolizumab RWD arm. PD-L1, programmed death-ligand 1; RWD, real-world data; TC, tumor cell.

**Figure S2.**
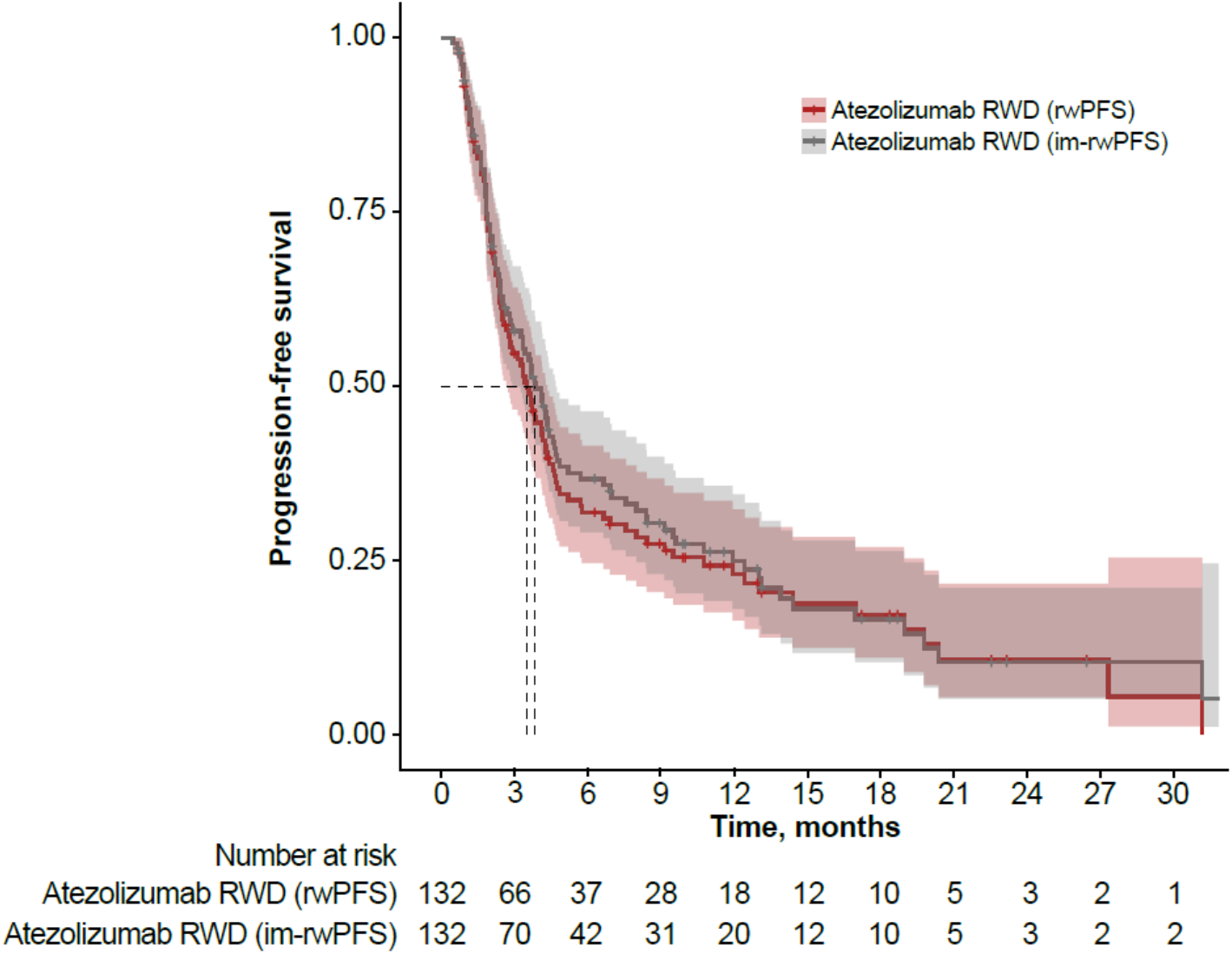
KM curves of atezolizumab rwPFS vs im-rwPFS. im-rwPFS, immune-modified-real-world progression-free survival; KM, Kaplan Meier; rwPFS, real-world progression-free survival.

**Figure S3.**
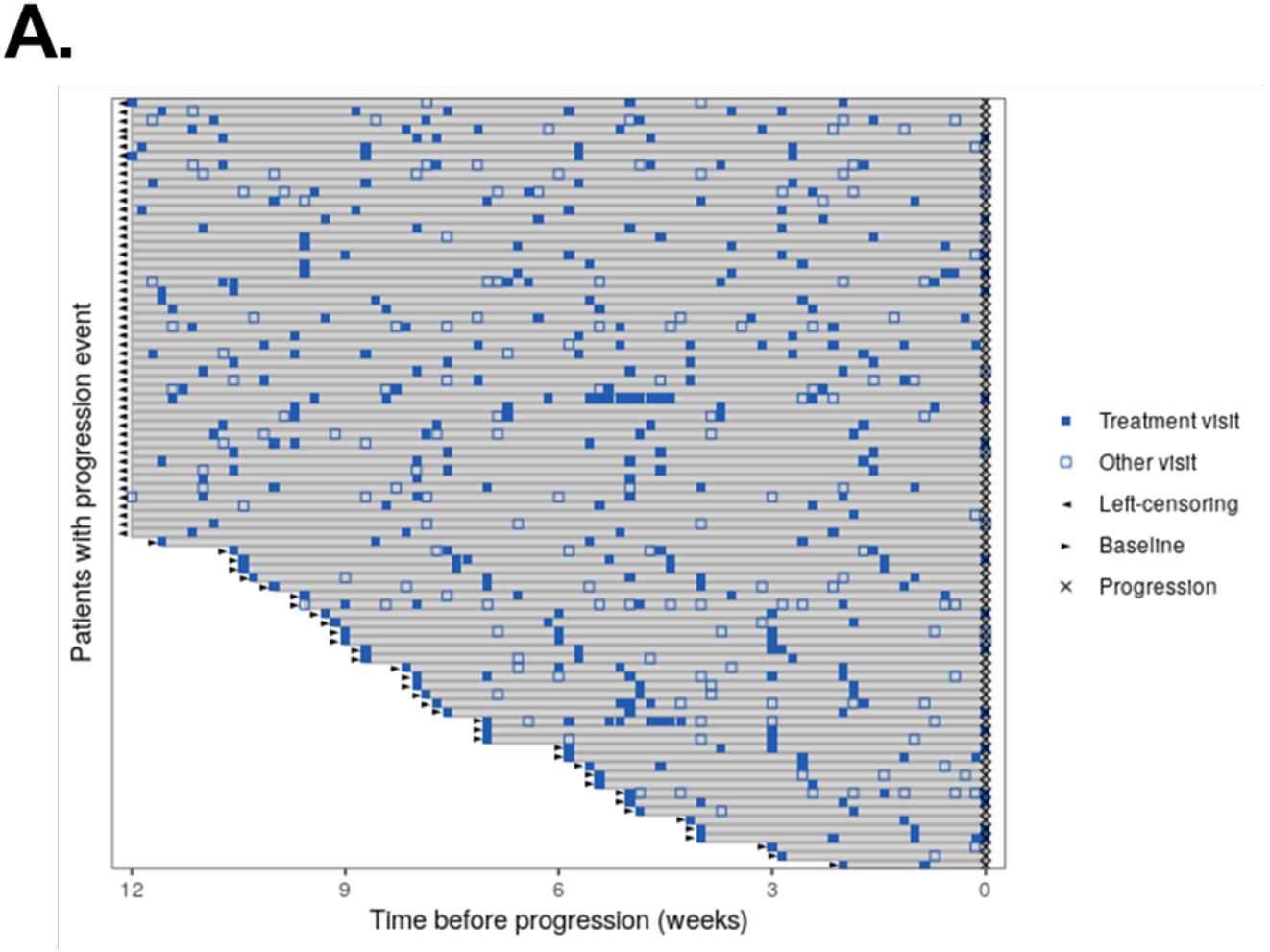
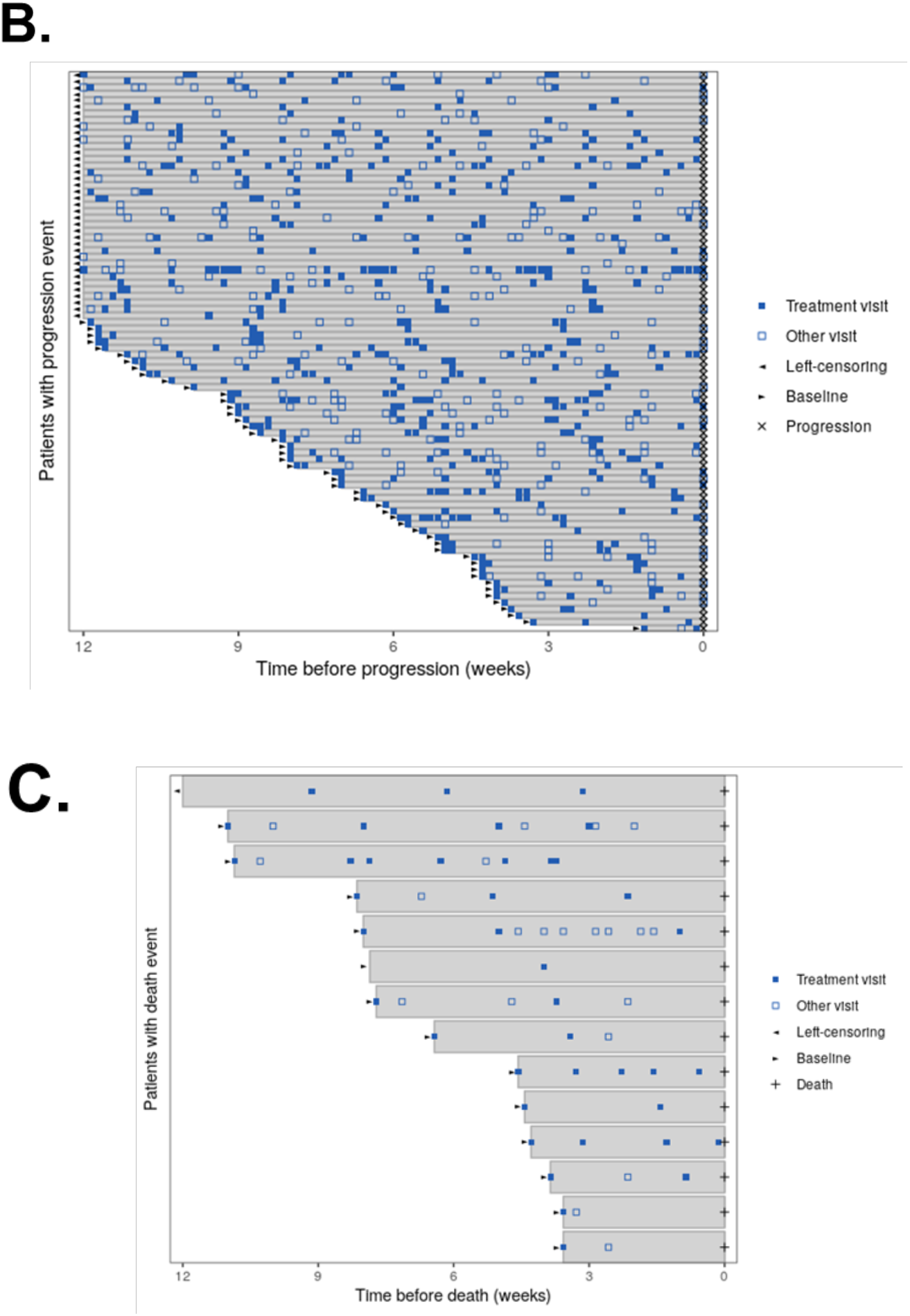
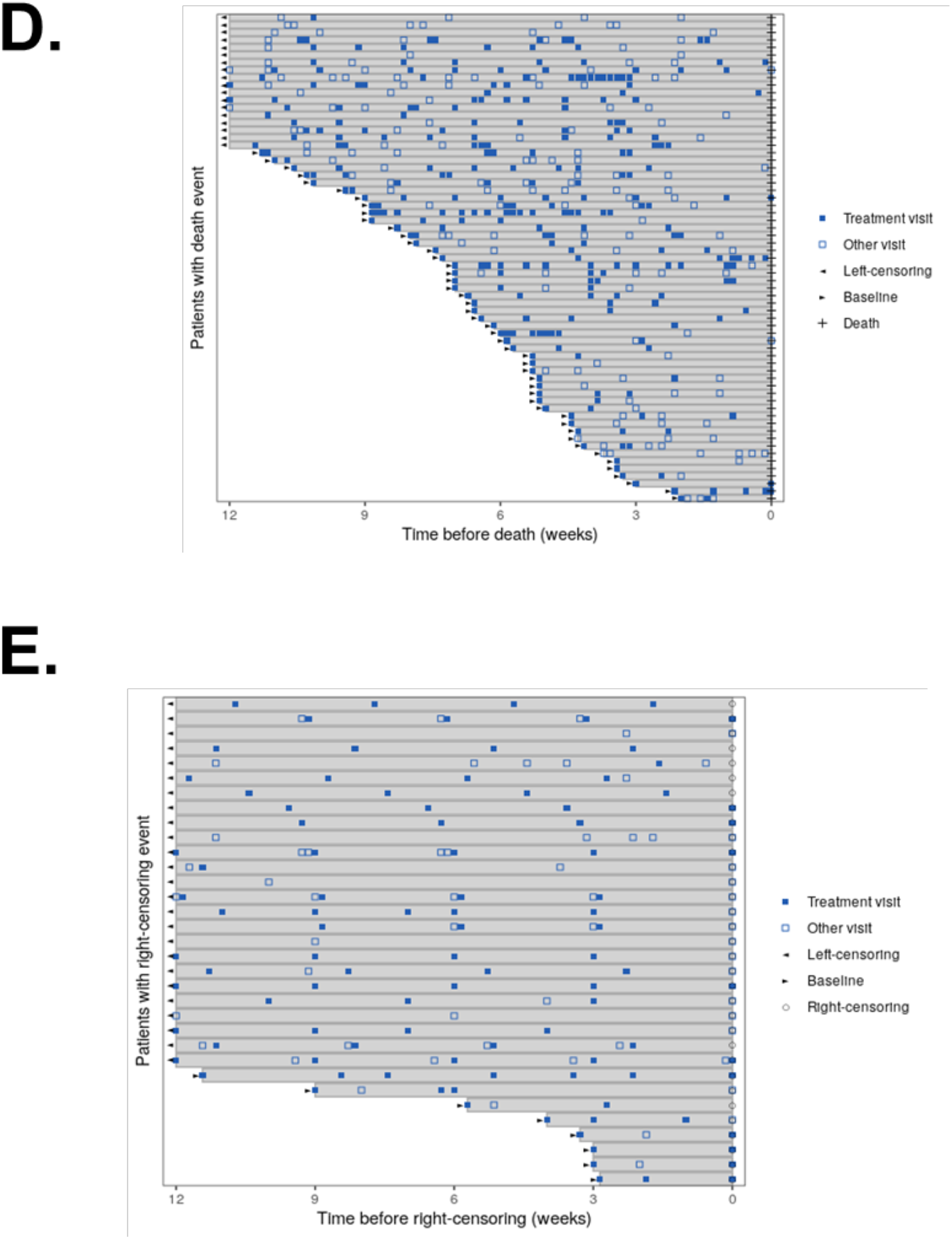
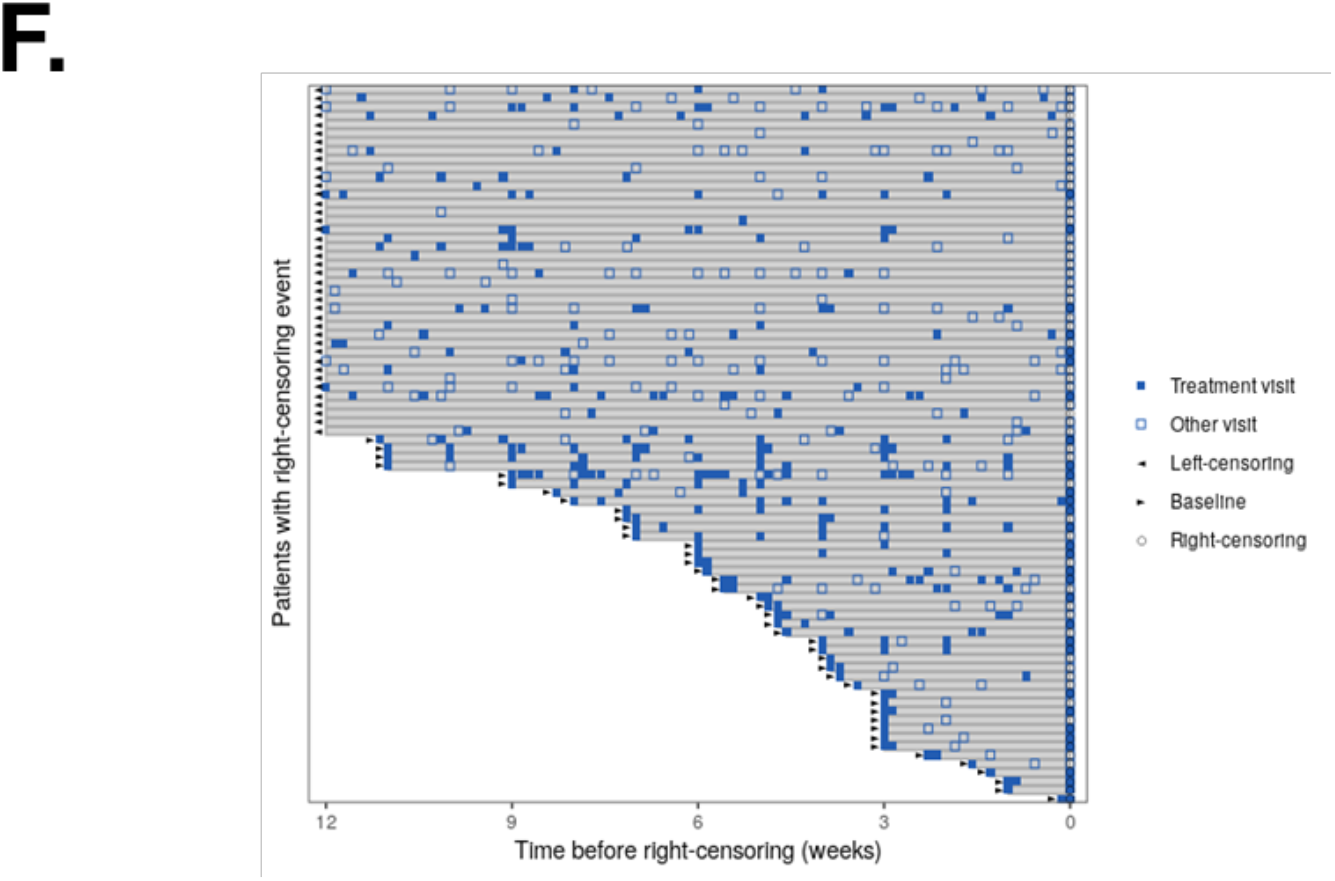
Frequency of patient visits for patients with a progression event in the A. atezolizumab arm and the B. docetaxel arm. Frequency of patient visits for patients with a death event in the C. atezolizumab arm or the D. docetaxel arm or a censoring event in the E. atezolizumab arm or F. docetaxel arm.

